# Barriers to Optimal Clinician Guideline Adherence in the Management of Markedly Elevated Blood Pressure: A Qualitative Content Analysis of Electronic Health Records

**DOI:** 10.1101/2024.01.12.24301223

**Authors:** Yuan Lu, Oreoluwa Arowojolu, Xiaoliang Qiu, Yuntian Liu, Leslie Curry, Harlan M. Krumholz

**Author notes:** **Corresponding Author:** Harlan M. Krumholz, MD, SM, 195 Church St, 5^th^ Floor, New Haven, CT, 06510. Co-First Authors (Yuan Lu, Oreoluwa Arowojolu).

## Abstract

**IMPORTANCE:** Hypertension poses a significant public health challenge. Despite clinical practice guidelines for hypertension management, clinician adherence to these guidelines remains suboptimal.

**OBJECTIVE:** This study aims to develop a taxonomy of suboptimal adherence scenarios for severe hypertension and identify barriers to guideline adherence.

**DESIGN:** We conducted a qualitative content analysis using electronic health records (EHRs) of Yale New Haven Health System who had at least two consecutive visits between January 1, 2013, and October 31, 2018.

**SETTING:** This was a thematic analysis of EHR data to generate a real-world taxonomy of scenarios of suboptimal clinician guideline adherence in the management of severe hypertension.

**PARTICIPANTS:** We identified patients with markedly elevated blood pressure ([BP]; defined as at least 2 consecutive readings of BP ≥160/100 mmHg) and no prescription for antihypertensive medication within a 90-day of the 2^nd^ BP elevation (n=4,828). We randomly selected 100 records from the group of all eligible patients for qualitative analysis.

**MAIN OUTCOMES AND MEASURES:** The scenarios and influencing factors contributing to clinician non-adherence to the guidelines for hypertension management.

**RESULTS:** Thematic saturation was reached after analyzing 100 patient records. Three content domains emerged: clinician-related scenarios (neglect and diffusion of responsibility), patient-related scenarios (patient non-adherence and patient preference), and clinical complexity-related scenarios (diagnostic uncertainty, maintenance of current intervention and competing medical priorities). Through a metareview of literature, we identified several plausible influencing factors, including a lack of protocols and processes that clearly define the roles within the institution to implement guidelines, infrastructure limitations, and clinicians’ lack of autonomy and authority, excessive workload, time constraints, clinician belief that intervention was not part of their role, or perception that guidelines restrict clinical judgment.

**CONCLUSIONS AND RELEVANCE:** This study illuminates reasons for suboptimal adherence to guidelines for managing markedly elevated BP. The taxonomy of suboptimal adherence scenarios, derived from real-world EHR data, is pragmatic and provides a basis for developing targeted interventions to improve clinician guideline adherence and patient outcomes.

**Key Points:** *Question:* What are the plausible scenarios and influencing factors contributing to clinician non-adherence to the guidelines for hypertension management?

*Findings:* In this qualitative study, we developed three domains of suboptimal adherence: clinician-related scenarios, patient-related scenarios, and clinical complexity-related scenarios; and identified several plausible influencing factors, including a lack of clear protocols and processes to implement guidelines, infrastructure limitations, and clinicians’ lack of autonomy and authority, excessive workload, time constraints, clinician belief or perception.

*Meaning:* This study introduces a taxonomy poised to inform targeted interventions, thereby enhancing guideline adherence and elevating care quality for severe hypertension.

## BACKGROUND

Hypertension, a chronic condition characterized by elevated blood pressure (BP), is a major public health concern affecting almost half of the US adult population. Patients with severely elevated blood pressure, defined as at least 2 consecutive readings of systolic BP ≥160 mmHg or diastolic BP ≥100 mm Hg, make up about 12% of all hypertensive patients and face an increased risk of complications,^1,2^ including severe and rapid systemic end-organ damage compared with those who have modestly elevated BP, thus requiring prompt and appropriate pharmacological treatment.^3^ The 2017 American College of Cardiology/American Heart Association guidelines recommend prompt evaluation and drug treatment followed by careful monitoring and upward dose adjustment in patients with severe hypertension.^3^ However, despite well-established clinical practice guidelines for the management of hypertension, adherence to these guidelines by clinicians remains suboptimal. A recent study based on electronic health record (EHR) data in the ambulatory setting found that almost 30% of patients with severely elevated BP had no active antihypertensive drug prescription before their second visit, and only 54% of those who were prescribed at least one antihypertensive drug class were prescribed the guideline-recommended two-drug class combination therapy.^4^ This finding highlights a missed opportunity to improve guideline adherence in this population.

Clinicians’ adherence to medication guidelines is a complex and multifaceted process that significantly impacts the implementation of evidence-based practice.^5^ The literature highlights various scenarios resulting in non-adherence to medication guidelines. These scenarios include situations where the recorded blood pressure does not accurately reflect the patient’s typical blood pressure, such as when home BPs are below the target range or when the patient is experiencing pain.^6^ In addition, scenarios such as the prioritization of other clinical concerns over hypertension, the need for ongoing monitoring and lifestyle counseling, and disagreements with specific recommendations also result in non-adherence.^6^ Moreover, how clinicians address patient-level factors, such as medication non-adherence and individual patient preferences, significantly influences guideline adherence. Clinician-level factors, including the belief that hypertension management is another clinician’s responsibility, further impact guideline adherence. Medication-related issues, like adverse drug events, use of medications from external sources, and contraindications, present additional adherence challenges.^6^ By recognizing and addressing these multifaceted factors, healthcare systems can implement strategies to improve clinician adherence to medication guidelines and enhance patient outcomes.

However, the current body of research on clinician guideline adherence in managing markedly elevated BP lacks a comprehensive identification of the reasons behind the inadequate treatment, particularly those based on routinely collected information during clinical practice, such as data from medical records. This information is particularly crucial as pharmacological interventions are vital in reducing BP and associated complications for this patient population. Furthermore, previous studies may have inadequately reported or underrepresented barriers to clinician guideline adherence, potentially due to methodological limitations.^5^ Consequently, we aimed to address these gaps by conducting a content analysis of EHRs to develop a comprehensive taxonomy of scenarios representing suboptimal guideline adherence in the ambulatory management of severe hypertension. This approach allowed us to identify plausible influencing factors contributing to clinician non-adherence. This information can potentially guide the creation and implementation of focused interventions, enhancing adherence to guidelines and quality of care for severe hypertension. Moreover, since the information is derived from real-world EHR data, it is pragmatic and enables the development of practical, automated EHR-based clinical decision support tools.^7^

## METHODS

### Data Sources

The data set consisted of data from adult patients at Yale New Haven Health System (YNHHS) who had at least two consecutive outpatient visits between January 1, 2013, and October 31, 2018. YNHHS is a large academic health system comprising five distinct hospitals and their associated ambulatory clinics in Connecticut and Rhode Island. The system provides services for approximately two million patients annually. All YNHHS hospitals used a secure, centralized EHR system designed by Epic Corporation to collect and store clinical and administrative data. The EHR data are maintained in a data repository at the YNHHS server. This study was approved by the institutional review board at Yale University and the need for informed consent was waived.

### Study Population

Eligible patients were 18 to 85 years old and had markedly elevated BP, defined as having measurements of systolic BP ≥160 mmHg or diastolic BP≥100 mmHg in at least 2 consecutive outpatient visits between January 1^st^, 2013 and October 31^st^, 2018, with no new antihypertensive medication prescription within 90 days of the index date. The index date was defined as the date of the 2^nd^ severely elevated BP reading. Patients with markedly elevated BP were selected as a focus given that the need to urgently achieve BP control in this population is unequivocal. Any two consecutive visits were required to be at least one day apart. We had access to all available data in the medical records, including patient demographics, past medical histories, vital signs, outpatient medications, laboratory results, encounter notes and scanned documents. A total of 4,828 patients met the eligibility criteria. We randomly selected 100 records from the group of all eligible patients for qualitative analysis, intending to select more if we did not achieve saturation (where no new concepts emerged from analyses of subsequent data).^8^

### Approach to Thematic Analysis and Taxonomy Development

Using a previously published inductive, systematic approach,^9–13^ we conducted a thematic analysis of EHR data to generate a real-world taxonomy of suboptimal clinician guideline adherence scenarios in managing severe hypertension. Then, we postulated plausible reasons/influencing factors/root causes for each taxonomized scenario based on contexts adopted from a previously published metareview of 25 systematic reviews on the factors influencing the implementation of clinical practice guidelines.^5^ We looked to extant literature because the provider encounter notes in the EHR often lacked comprehensive information regarding the reasons/influencing factors/root causes for a clinician’s decision to not initiate or intensify treatment in patients with markedly elevated BP.

#### Step 1: Development of Rubric for Medical Chart Review

Through an iterative process, a team of three clinicians and/or experienced cardiovascular researchers (O.A., Y.L., H.K.) developed a rubric to systematically abstract data from the EHR. We obtained demographic data (including age, sex, race, and ethnicity) and clinical data relevant to the diagnosis and treatment of hypertension (including BP measurements, medical history, medication prescriptions, and medical context of the encounter) and established criteria for consistency (to support explicit review). Additionally, the data extraction rubric was designed to offer flexibility, allowing reviewers to go beyond strict numerical or binary criteria and make subjective assessments. This approach included evaluating the rationale behind a clinician’s decisions, considering the medical context of each encounter, and interpreting data points with a nuanced understanding of patient history, comorbidities, or unique clinical scenarios.

Furthermore, while the rubric establishes consistency criteria, it also provides guidance for implicit review, enabling reviewers to use their clinical judgment to uncover underlying reasons for suboptimal adherence to guidelines not explicitly stated in the EHR (implicit review).^13–16^

#### Step 2: Data Abstraction

Two abstractors (O.A., X.Q.) participated in a training session, during which they collectively abstracted 15 medical records using the rubric described above and generated a narrative summary for each case. Decision rules and operational definitions were refined to reduce ambiguity and to facilitate standardized data abstraction. Discrepancies were resolved during face-to-face meetings with discussion among all reviewers until consensus was reached. Once the rubric was finalized, each abstractor reviewed a random sample of 50 medical records respectively. In the end, 100 cases were reviewed when reviewers determined they reached saturation; that is, no new constructs emerged from reviewing subsequent cases.^8^

#### Step 3: Content Analysis and Taxonomy Development

The 100 cases abstracted using the rubric above were analyzed using conventional content analysis. Content analysis is a systematic, replicable technique for compressing many words of text into fewer content categories based on explicit coding rules.^17,18^ Content analysis enables researchers to sift through large volumes of data with relative ease in a systematic fashion and it is useful in examining the patterns in documentation.^19^

We used emergent coding and established categories following a preliminary examination of the abstracted data obtained in Step 2. First, one author (O.A.) independently reviewed the abstracted data and identified a set of suboptimal clinician guideline adherence scenarios to form the initial code list, which was then developed into a consolidated code book. Second, two authors (O.A., Y.L.) reviewed this code book for face validity and revised it based on group discussion. Third, the consolidated code book was trialed on 10 cases by the coding group (O.A., Y.L.) to ensure consistent coding application. The coding group checked that the reliability of the coding was established (agreement >95%). Then all 100 cases were coded by the coding group. Finally, a larger author group (O.A., Y.L., L.C., H.M.K.) used an iterative, consensus-based discussion process to group the coding into major content themes with sub-themes, maintaining a consensus and primary data referencing approach.^20^

#### Step 4: Linking taxonomized scenarios with influencing factors

Members of the author group (O.A., Y.L., L.C., H.M.K.), are experienced clinicians and cardiovascular researchers engaged in a detailed and collaborative process to identify influencing factors for each categorized scenario. This process began with drafting initial hypotheses based on their expertise and insights. These drafts were then circulated among the group for review. Each member provided their feedback and perspectives, leading to a series of discussions. Through these iterative discussions, the group refined their ideas, ensuring a robust consensus was reached on the plausible influencing factors. This consensus was grounded in the contexts adopted from the previously published metareview of 25 systematic reviews on factors influencing the implementation of clinical practice guidelines.^5^

## RESULTS

### Study Sample Characteristics

We generated a randomized list of 200 patients and reached saturation with 100 patients. The mean age at the index visit was 66.5 (standard deviation [SD], 12.8) years; 50% were female; 85%, 8%, and 5% were noted in the EHR to be White, Black, and Latino/Hispanic, respectively. A total of 31% had private insurance, 58% had Medicare, 11% had Medicaid and 0% did not have health insurance. The mean (SD) SBP and DBP of the sample at the index date was 166.2 (11.5) mmHg and 87.7 (12.7) mmHg, respectively. The median (interquartile range) time between visits was 42 (18 to 85) days. A large proportion of patients had comorbidities at the index date, including 44% patients with obesity (BMI≥ 30kg/m^2^), 16% with diabetes, 5% with chronic kidney disease, 36% with cancer.

### Content Domains

Based on a thematic analysis of data available in the EHR for patients meeting our criteria, we identified a variety of scenarios of suboptimal clinician guideline adherence in managing severe hypertension pertaining to either non-initiation or non-intensification of pharmacological therapy (**Table 1**). Non-initiation of pharmacological treatment was defined as an absence of the initiation of antihypertensive therapy in response to severely elevated BP in a patient with at least 2 consecutive readings of severely elevated BP. Non-intensification of pharmacological treatment was defined as failure to intensify/modify treatment or initiate an urgent referral on the index visit for a patient with severely elevated BP who was previously on antihypertensives.

**Table 1.** Scenarios of suboptimal clinician guideline adherence in the management of severe hypertension identified under non-initiation or non-intensification of anti-hypertensive treatment (order of frequency from most to least)

| **Non-initiation of treatment** | **Non-intensification of treatment** |
| --- | --- |
| Neglect | Neglect |
| Patient preference | Diffusion of responsibility |
| Diffusion of responsibility | Patient non-adherence |
| Diagnostic uncertainty with BP measurement | Patient preference |
|  | Diagnostic uncertainty with BP measurement |
|  | Maintenance of current BP intervention |
|  | Competing medical priorities |

These identified scenarios (subcategories) of suboptimal clinician guideline adherence were taxonomized and grouped into 3 main content domains: clinician-related scenarios, patient-related scenarios, and clinical complexity-related scenarios (**Tables 2**). **Tables 3a-c** include example quotations or clinical situations pertaining to each scenario.

#### Clinician-related Scenarios

Clinician-related scenarios were defined as instances where clinicians did not initiate or intensify antihypertensive treatment due to factors related to their intentions, capabilities, or scope. Under this main content domain, we identified 2 subcategories: neglect and diffusion of responsibility. Neglect included instances in which the clinician encountered on the index date did not acknowledge nor prioritize the BP at the visit. For example, **Table 3a** highlights a clinical scenario in which a patient who presented to a provider for wound care had a second consecutive markedly elevated BP reading at presentation, but this was not addressed in the encounter note, nor was any action or intervention relating to the BP carried out. Diffusion of responsibility included instances in which the specialist visited did not initiate/intensify treatment, explicitly displacing responsibility to a hypertension-managing provider (i.e., PCP, cardiology, etc.), excluding cases where an urgent referral to the provider was made.

**Table 2.** Comprehensive codebook of scenarios of suboptimal clinician guideline adherence in the management of severe hypertension.

| Main Content Domain | Subcategory | Description |
| --- | --- | --- |
| Clinician-related scenarios |  | Scenarios of non-initiation/non-intensification of antihypertensive treatment by the clinician due to issues relating to clinician intention, capability or scope. |
|  | Neglect | Clinician did not acknowledge nor prioritize the BP at visit |
|  | Diffusion of responsibility | Specialist visited did not initiate/intensify treatment, opting instead to defer responsibility to a hypertension-managing provider (i.e., PCP, cardiologist, etc.), excluding cases where an urgent referral to said provider was made. |
| Patient-related scenarios |  | Scenarios of non-initiation/non-intensification of antihypertensive treatment by the clinician due to patient behavioral considerations. |
|  | Patient non-adherence | Clinician did not intensify intervention due to patient's non-adherence to current therapy. |
|  | Patient preference | Clinician did not initiate/intensify intervention due to patient preference not to. |
| Clinical complexity-related scenarios |  | Scenarios of non-initiation/non-intensification of antihypertensive treatment by the clinician due to clinical situational complexities. |
|  | Diagnostic uncertainty with BP measurement | The clinician did not initiate or intensify treatment due to variations in BP measurements, whether at home or in the office. This includes cases where high in-office readings were believed to be a result of the white coat effect, pain, or other causes of reactive elevated BP. |
|  | Maintenance of current BP intervention | Clinician chose to delay intensifying treatment in order to observe if current antihypertensives/lifestyle modifications would result in BP control. |
|  | Competing medical priorities | Clinician did not intensify treatment due to competing clinical considerations |

Abbreviation: BP: blood pressure; PCP: primary care provider.

**Table 3a.**
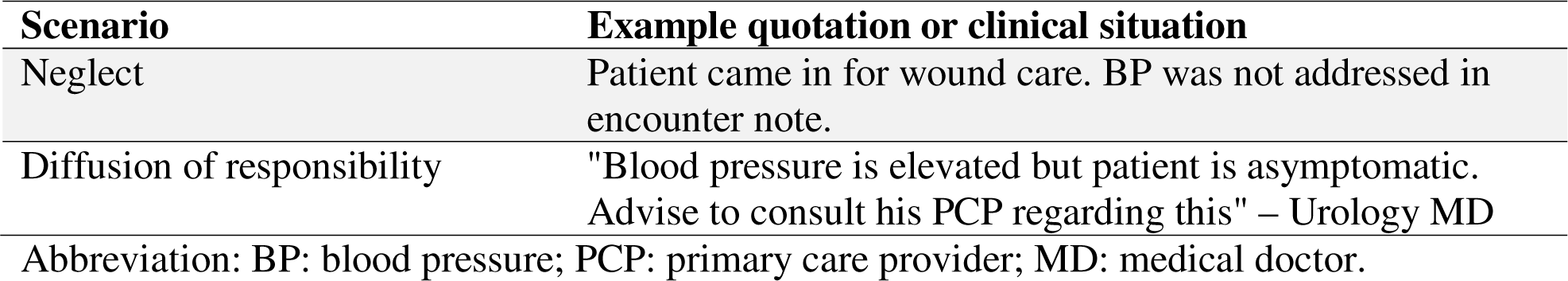
Example quotations and clinical situations illustrating clinician-related scenarios.

#### Patient-related Scenarios

Patient-related scenarios were defined as instances where clinicians did not initiate or intensify antihypertensive treatment due to considerations related to patient behavior. Under this main content domain, we identified 2 subcategories: patient non-adherence and preference. Patient non-adherence included instances where the clinician did not intensify intervention due to the patient’s non-adherence to current therapy. For example, our analysis identified a case in which a patient who had previously had adequate BP control on metoprolol had not taken his medication in two days when he presented with markedly elevated BP and the clinician decided to counsel the patient on adherence rather than modify or intensify treatment at the visit (**Table 3b**). Patient preference included instances where the clinician did not initiate nor intensify intervention due to patient preference.

**Table 3b.** Example quotations and clinical situations illustrating patient-related scenarios.

| **Scenario** | **Example quotation or clinical situation** |
| --- | --- |
| Patient non-adherence | "Patient noted to take metoprolol, however he has not taken his medications for two days...this would explain his tachycardia and increase in NIBP measurement." |
| Patient preference | "Patient states that he has some element of white coat hypertension and that his blood pressure tends to run lower. Does not want medication at this time." |
| Abbreviation: BP: blood pressure; NIBP: non-invasive blood pressure. |  |

**Table 3c.** Example quotations and clinical situations illustrating clinical complexity-related scenarios.

| **Scenario** | **Example quotation or clinical situation** |
| --- | --- |
| Diagnostic uncertainty with BP measurement | "BP on arrival 170/98 mmHg. Patient states she was advised to follow up with PCP after appointments at cancer center in February. Patient stated she did, but that her blood pressure is only elevated when she is at cancer center." |
| Maintenance of current BP intervention | "Continue meds. Continue BP check at home." |
| Competing medical priorities | "We would recommend no treatment at this time...especially since she has significant history of kidney injury." |

Abbreviation: BP: blood pressure; PCP: primary care provider.

#### Clinical complexity-related Scenarios

These scenarios involve instances where clinicians did not initiate or intensify antihypertensive treatment due to the complexities of the clinical situation. Under this main content domain, we identified 3 subcategories, namely diagnostic uncertainty with BP measurement, maintenance of current BP intervention, and competing medical priorities. Diagnostic uncertainty with BP measurement included cases where the clinician did not initiate/intensify treatment due to variation in BP measurements, either at home or in the office, including cases where high in-office readings were thought to be due to white coat effect or pain. Maintenance of current BP intervention included cases where the clinician chose to delay intensifying treatment to observe if current antihypertensives and/or lifestyle modifications would result in BP control. Competing medical priorities included cases in which the clinician chose to delay intensifying treatment due to several competing medical conditions.

### Barriers to Optimal Clinician Guideline Adherence in the Management of Markedly Elevated Blood Pressure

For each scenario of suboptimal clinician guideline adherence in the management of markedly elevated BP, we hypothesized relevant plausible influencing factors or root causes, which are barriers to optimal guideline adherence. These factors were identified based on barriers frequently reported in the literature^6^ within the following contexts: health organization, health professional, patient, and guideline (**Table 4**).

**Table 4.** Plausible influencing factors for scenarios of suboptimal clinician guideline medication adherence, based on analysis of metareview findings.

| Scenario | Plausible influencing factors |
| --- | --- |
| Neglect | **Health organization context**<br>• Absence of a leader that establishes priorities<br>• Lack of protocols and processes that clearly define the roles within the institution to implement guidelines<br>• Too little time in the medical consultations<br>• Excessive workload<br>• Deficiency in staff continuous education<br>• Deficiencies in the referral of patients to services<br>• Lack of skill and specialist knowledge within services<br>• High turnover of staff that prevents a continuous training process<br>• Lack of coordination and disagreement among staff<br>• Financial constrains for the adoption of new interventions<br>• Lack of availability of interpreters in services<br>• Lack of access to information, lack of mechanisms and systems to support storing of information<br>**Health professional context**<br>• Lack of effective communication, research, and self-learning skills<br>• Little familiarity with guideline recommendations<br>• Lack of autonomy and authority<br>• Belief that intervention was not part of their role<br>**Guideline context**<br>• Lack of clarity of guidelines |
| Diffusion of responsibility | **Health organization context**<br>• Lack of protocols and processes that clearly define the roles within the institution to implement guidelines<br>• Too little time in the medical consultations<br>• Excessive workload<br>• Deficiency in staff continuous education<br>• Deficiencies in the referral of patients to services<br>• Lack of skill and specialist knowledge within services<br>• Insufficient support from institutions<br>• High turnover of staff that prevents a continuous training process<br>• Limitations of infrastructure |
|  | • Lack of coordination and disagreement among staff<br>• Lack of availability of interpreters in services<br>• Lack of access to information, lack of mechanisms and systems to support storing of information<br>**Health professional context**<br>• Lack of effective communication, research, and self-learning skills<br>• Little familiarity with guideline recommendations<br>• Lack of autonomy and authority<br>• Belief that intervention was not part of their role<br>**Guideline context**<br>• Lack of clarity of guidelines |
| Patient non-adherence & Patient Preference | **Health professional context**<br>• Physician's reluctance to use guidelines because of patient factors, self-belief, or fear of complications<br>**Patient context**<br>• Lack of motivation, compliance, and knowledge to follow the recommendations<br>• Unawareness about the health organization characteristics and their disease<br>• Patients' financial situation and occupational status<br>• Depression, anxiety, and fear<br>**Guideline context**<br>• Beliefs that guidelines are too rigid, may not always be practical and cannot be applied on a day-to-day |
| Diagnostic uncertainty with BP measurement | **Health organization context**<br>• Lack of access to information, lack of mechanisms and systems to support storing of information<br>**Guideline context**<br>• Beliefs that guidelines are too rigid, may not always be practical and cannot be applied on a day-to-day<br>• Guidelines restrict clinical judgment and challenge professional autonomy and limits treatment options |
| Maintenance of current BP intervention | **Health professional context**<br>• Greater confidence in clinical experience than in guidelines recommendations<br>• Physician's reluctance to use guidelines because of patient factors, self-belief, or fear of complications<br>**Patient context**<br>• Patient comorbidities, mobility problems, polypharmacy, and self-efficacy<br>**Guideline context**<br>• Lack of awareness of the existence of guidelines and |
|  | clarity of guidelines<br>Guidelines restrict clinical judgment and challenge professional autonomy and limits treatment options |
| Competing medical priorities | **Health organization context**<br>Absence of a leader that establishes priorities<br>**Patient context**<br>Patient comorbidities, mobility problems, polypharmacy, and self-efficacy |
| Synthesized from Correa et al.<sup>6</sup> |  |

Clinician-related scenarios, specifically neglect and diffusion of responsibility, may be influenced by various factors within the health organization context. These factors could include a lack of protocols and processes that clearly define the roles within the institution to implement guidelines, too little time in the medical consultations, excessive workload, or infrastructure limitations. Within the health professional context, factors such as lack of autonomy and authority or the belief that intervention was not part of their role may contribute to these scenarios. Additionally, guideline factors, such as lack of clarity of guidelines, can play a role.

Patient-related scenarios, including patient non-adherence and preference, may arise due to several factors within different contexts. Within the health professional context, clinician reluctance to use guidelines due to patient factors, self-belief, or fear of complications may influence these scenarios. Patient factors, such as unawareness of their conditions or a lack of motivation, compliance, and knowledge to follow recommendations can also contribute. Furthermore, guideline factors, such as beliefs that guidelines are too rigid and may not always be practical in certain patient contexts may play a role.

Clinical complexity-related scenarios, encompassing diagnostic uncertainty with BP measurement, maintenance of current intervention, and competing medical priorities, can be influenced by factors within multiple contexts. Within the health organization context, factors such as lack of access to information, and lack of mechanisms and systems to support information storage may contribute. In the health professional context, greater confidence in clinical experience compared to guidelines recommendations can be a factor. Patient factors, such as comorbidities, mobility limitations, polypharmacy, and self-efficacy, can also play a role. Finally, guideline factors, such as the perception that guidelines restrict clinical judgment, challenge professional autonomy, and limit treatment options, can contribute to these scenarios.

## DISCUSSION

This study provides novel insights into the factors contributing to suboptimal adherence to guidelines among clinicians treating patients with markedly elevated blood pressure in ambulatory settings. Our taxonomy, derived from real-world EHR data, not only categorizes these instances but also describes the factors influencing each scenario of suboptimal adherence. Such a pragmatic framework is poised to inform targeted interventions, thus enhancing adherence and patient outcomes.

Our study advances the existing body of literature in several ways. We have previously detailed various mechanisms through which patients experience persistent hypertension, such as the lack of intensification in pharmacological treatment, failure to implement prescribed therapies, and non-response to treatment.^10^ Building on this, the current study specifically illuminates the mechanisms behind clinicians’ failure to treat ambulatory patients with severely elevated BP effectively and explores the reasons for these shortcomings. To our knowledge, this is the first study to develop a taxonomy for categorizing instances of suboptimal clinician adherence to guidelines in managing patients with markedly elevated BP using real-world data. It also pioneers in identifying contributing factors at the organizational, professional, patient, and guideline levels. Compared with prior work on clinician guideline adherence, a key strength of this study is its foundation in EHR data. EHRs capture a broad spectrum of real-world clinical interactions across diverse patient demographics, enhancing our findings’ practicality and external validity.^7^ Research based on EHR data can inform more effective clinical decisions by evaluating the quality and cost implications of guideline-conformant care for chronic conditions such as hypertension. Furthermore, EHRs assist in pinpointing the issue of suboptimal clinician guideline adherence in the management of significantly elevated BP and serve as a robust framework for integrating potential solutions.

Our analysis has identified not only instances of non-intensification but also numerous cases of non-initiation of antihypertensive treatment in patients with markedly elevated BP. This finding is particularly troubling considering that a BP reading of ≥160/100 mmHg in a patient not on antihypertensive medication would typically warrant immediate clinical action. The clinician-related scenarios we have identified, including neglect and diffusion of responsibility, are increasingly concerning within today’s healthcare environment.^21,22^ Neglect often occurs when a patient’s primary reason for visiting the healthcare provider is unrelated to hypertension. Meanwhile, diffusion of responsibility arises when clinicians may defer action because they believe someone else, possibly more specialized, should take the lead^23^ —this is evidenced in cases where care providers defer the implementation of guidelines to hypertension specialists, even when faced with significantly elevated BP readings. The willingness of clinicians to adhere to guidelines may also be hindered by patient preferences or issues with patient adherence to therapy, which are well-documented challenges.^24–26^ Additionally, our study confirms that adherence is affected by clinical complexities, such as ambiguity in BP measurements and competing health conditions, echoing the findings of other research in the field.

Our analysis underscores the necessity of addressing the multi-dimensional nature of guideline non-adherence. Under the proposed taxonomy, each category and subcategory of non-adherence scenarios is linked to specific causes and targeted interventions. For example, scenarios affected by organizational factors may improve with robust leadership, clear priorities, sufficient staffing, knowledge-sharing forums, streamlined processes, and regular, communicative audits with constructive feedback. ^5,25,27,28^ Health organizations can further support clinician adherence by integrating evidence-based decision support tools within EHR systems, such as automated alerts, reminders, and advanced patient portals, along with improved collaborative tools for care teams.^29^ Addressing health professional-level causes involves fostering a willingness to embrace new practices, educating about guidelines, and reinforcing personal accountability.^7,30^ For patient-level factors, strategies include raising health awareness, early education, clear communication about the impact of non-adherence, and peer support. Concerning the guidelines themselves, simplifying their presentation, tailoring them to the local context, and involving end-users in their development can enhance their usability and adherence.^7,24^

Reflecting on the broader implications, this study’s findings can stimulate healthcare policies aimed at systematizing adherence to guidelines and thus improve the quality of care delivered. This is particularly pertinent in light of our identification of implicit bias and structural racism as underlying factors contributing to non-adherence, which are critical to address in the pursuit of equitable healthcare.^31,32^

While our study’s EHR-based nature significantly enhances its applicability, there are several limitations. First, this study employed qualitative analysis of EHR data, which does not yield detailed descriptive statistics regarding instances of non-adherence to clinical guideline at our institution. Secondly, encounter notes within the EHR may not always provide sufficient detail to conclusively ascertain the intentions or rationale underpinning specific clinical decisions. Additionally, the study’s reliance on the reviewers’ judgment, coupled with the breadth and quality of the referenced meta-review,^6^ could potentially influence the determination of factors contributing to the identified scenarios of non-adherence.

## CONCLUSION

In conclusion, by highlighting the multifaceted reasons for suboptimal guideline adherence, our study provides a foundation for developing nuanced interventions. As we look towards a future of healthcare that is both evidence-based and patient-centered, it is imperative that we consider the complex interplay of factors at the organizational, professional, patient, and guideline levels that influence clinician behaviors.

## Data Availability

All data produced in the present study are available upon reasonable request to the authors.

## ACKNOWLEDGMENT

Drs. Yuan Lu and Harlan Krumholz had full access to all the data in the study and takes responsibility for the integrity of the data and the accuracy of the data analysis.

## Data Sharing Statement

All supporting data of this study are available upon reasonable request from the corresponding author.

## REFERENCES

1. Muntner P, Hardy ST, Fine LJ, et al. Trends in Blood Pressure Control Among US Adults With Hypertension, 1999-2000 to 2017-2018. JAMA. 2020;324(12):1190–1200. doi:10.1001/jama.2020.14545

2. Lu Y, Huang C, Mahajan S, et al. Leveraging the Electronic Health Records for Population Health: A Case Study of Patients With Markedly Elevated Blood Pressure. J Am Heart Assoc. 2020;9(7):e015033. doi:10.1161/JAHA.119.015033

3. Whelton PK, Carey RM, Aronow WS, et al. 2017 ACC/AHA/AAPA/ABC/ACPM/AGS/APhA/ASH/ASPC/NMA/PCNA Guideline for the Prevention, Detection, Evaluation, and Management of High Blood Pressure in Adults: A Report of the American College of Cardiology/American Heart Association Task Force on Clinical Practice Guidelines. J Am Coll Cardiol. 2018;71(19):e127–e248. doi:10.1016/j.jacc.2017.11.006

4. Lu Y, Huang C, Liu Y, et al. Medication Guideline Adherence Among Patients with Markedly Elevated Blood Pressure in A Real-World Setting. medRxiv. 2022:2022.02. 16.22271094.

5. Correa VC, Lugo-Agudelo LH, Aguirre-Acevedo DC, et al. Individual, health system, and contextual barriers and facilitators for the implementation of clinical practice guidelines: a systematic metareview. Health Res Policy Syst. 2020;18(1):74. doi:10.1186/s12961-020-00588-8

6. Lin ND, Martins SB, Chan AS, et al. Identifying barriers to hypertension guideline adherence using clinician feedback at the point of care. AMIA Annu Symp Proc. 2006;2006:494–8.

7. Chan WV, Pearson TA, Bennett GC, et al. ACC/AHA Special Report: Clinical Practice Guideline Implementation Strategies: A Summary of Systematic Reviews by the NHLBI Implementation Science Work Group: A Report of the American College of Cardiology/American Heart Association Task Force on Clinical Practice Guidelines. J Am Coll Cardiol. 2017;69(8):1076–1092. doi:10.1016/j.jacc.2016.11.004

8. Patton MQ. Qualitative research & evaluation methods: Integrating theory and practice. Sage publications; 2014.

9. Curry LA, Nembhard IM, Bradley EH. Qualitative and mixed methods provide unique contributions to outcomes research. Circulation. 2009;119(10):1442–52. doi:10.1161/CIRCULATIONAHA.107.742775

10. Lu Y, Xinxin Du C, Khidir H, et al. Developing an Actionable Taxonomy of Persistent Hypertension Using Electronic Health Records. Circ Cardiovasc Qual Outcomes. 2023;16(3):e009453. doi:10.1161/CIRCOUTCOMES.122.009453

11. Butler CR, Wightman A, Richards CA, et al. Thematic Analysis of the Health Records of a National Sample of US Veterans With Advanced Kidney Disease Evaluated for Transplant. JAMA Intern Med. 2021;181(2):212–219. doi:10.1001/jamainternmed.2020.6388

12. O’Hare AM, Butler CR, Taylor JS, et al. Thematic Analysis of Hospice Mentions in the Health Records of Veterans with Advanced Kidney Disease. J Am Soc Nephrol. 2020;31(11):2667–2677. doi:10.1681/ASN.2020040473

13. Bradley EH, Curry LA, Devers KJ. Qualitative data analysis for health services research: developing taxonomy, themes, and theory. Health Serv Res. 2007;42(4):1758–72. doi:10.1111/j.1475-6773.2006.00684.x

14. Harrison ER, Kahn KL, Sherwood MJ, et al. Structured Implicit Review of the Medical Record: A Method for Measuring the Quality of Inhospital Medical Care and a Summary of Quality Changes Following Implementation of the Medicare Prospective Payment System. 1991;

15. Hutchinson A, Coster J, Cooper K, et al. Creating and designing the healthcare experience. The International Ergonomics Association. 2008;

16. Ashton CM, Kuykendall DH, Johnson ML, Wray NP. An empirical assessment of the validity of explicit and implicit process-of-care criteria for quality assessment. Med Care. 1999;37(8):798–808. doi:10.1097/00005650-199908000-00009

17. Krippendorff K. Content analysis: An introduction to its methodology. Sage publications; 2018.

18. Weber RP. Basic content analysis. vol 49. Sage; 1990.

19. Stemler S. An overview of content analysis. Practical Assessment. Research & Evaluation. 2001;7(17):137–146.

20. Dang DT, Nguyen NT, Hwang D. Multi-step consensus: an effective approach for determining consensus in large collectives. Cybernetics and Systems. 2019;50(2):208–229.

21. Marcotte LM, Krimmel-Morrison J, Liao JM. How to Keep Diffusion of Responsibility From Undermining Value-Based Care. AMA J Ethics. 2020;22(9):E802–807. doi:10.1001/amajethics.2020.802

22. Oyebode F. Clinical errors and medical negligence. Med Princ Pract. 2013;22(4):323–33. doi:10.1159/000346296

23. McIntosh E. The implications of diffusion of responsibility on patient safety during anaesthesia, ‘So that others may learn and even more may live’ - Martin Bromiley. J Perioper Pract. 2019;29(10):341–345. doi:10.1177/1750458918816572

24. Bierbaum M, Rapport F, Arnolda G, et al. Clinical practice guideline adherence in oncology: A qualitative study of insights from clinicians in Australia. PLoS One. 2022;17(12):e0279116. doi:10.1371/journal.pone.0279116

25. Kanazaki R, Smith B, Girgis A, Connor SJ. Clinician Adherence to Inflammatory Bowel Disease Guidelines: Results of a Qualitative Study of Barriers and Enablers. Crohns Colitis 360. 2023;5(3):otac018. doi:10.1093/crocol/otac018

26. Cardona M, Craig L, Jones M, et al. Guideline Adherence As An Indicator of the Extent of Antithrombotic Overuse and Underuse: A Systematic Review. Glob Heart. 2022;17(1):55. doi:10.5334/gh.1142

27. Lau R, Stevenson F, Ong BN, et al. Achieving change in primary care--causes of the evidence to practice gap: systematic reviews of reviews. Implement Sci. 2016;11:40. doi:10.1186/s13012-016-0396-4

28. Craig LE, McInnes E, Taylor N, et al. Identifying the barriers and enablers for a triage, treatment, and transfer clinical intervention to manage acute stroke patients in the emergency department: a systematic review using the theoretical domains framework (TDF). Implement Sci. 2016;11(1):157. doi:10.1186/s13012-016-0524-1

29. Lu Y, Melnick ER, Krumholz HM. Clinical decision support in cardiovascular medicine. BMJ. 2022;377:e059818. doi:10.1136/bmj-2020-059818

30. Khatib R, Schwalm JD, Yusuf S, et al. Patient and healthcare provider barriers to hypertension awareness, treatment and follow up: a systematic review and meta-analysis of qualitative and quantitative studies. PLoS One. 2014;9(1):e84238. doi:10.1371/journal.pone.0084238

31. Linnander EL, Ayedun A, Boatright D, et al. Mitigating structural racism to reduce inequities in sepsis outcomes: a mixed methods, longitudinal intervention study. BMC Health Serv Res. 2022;22(1):975. doi:10.1186/s12913-022-08331-5

32. Williams DR, Lawrence JA, Davis BA. Racism and Health: Evidence and Needed Research. Annu Rev Public Health. 2019;40:105–125. doi:10.1146/annurev-publhealth-040218-043750

